# Household transmission of COVID-19, Shenzhen, January-February 2020

**DOI:** 10.1101/2020.05.11.20092692

**Authors:** Lan Wei, Qiuying Lv, Ying Wen, Shuo Feng, Wei Gao, Zhigao Chen, Bin Cao, Yan Lu, Xiaoliang Wu, Jin Zhao, Xuan Zou, Tiejian Feng, Ben Cowling, Shujiang Mei

## Abstract

Coronavirus disease 2019 has led to more than three million cases globally. Since the first family cluster of COVID-19 cases identified in Shenzhen in early January, most of the local transmission occurred within household contacts. Identifying the factors associated with household transmission is of great importance to guide preventive measures.

## Texts

Coronavirus disease 2019 (COVID-19) caused by SARS-CoV-2 infection was first identified in Wuhan in December 2019. As of Apr 29, 2020, it has led to more than three million confirmed cases globally (1). China was able to bring its first wave of infections under control through the use of social distancing and a rapid expansion in laboratory testing capacity (2). More than one billion people were confined to their homes across China, including about 13 million people in the rapidly developing city of Shenzhen in southern China. Since the first family cluster of COVID-19 cases identified in Shenzhen in early January 2020, most of the local transmission occurred within household contacts (3), similar to the situation elsewhere in China (4). We analyzed the dynamics of household transmission in Shenzhen to identify the risk factors for transmission and the effectiveness of preventive measures.

## The Study

From Jan 1 to Feb 14, 2020 we identified 23 households in which there were a total of 139 individuals, including 60 confirmed COVID-19 cases and their 79 household contacts. Informed consent was waived as data collection was part of the continuing public health investigation of an emerging outbreak. This study was approved by the ethics committees of Shenzhen Center for Disease Control and Prevention. The mean age of the index cases was 33.9 years old (range 2 to 67), with 35(58.3%) of them were female. In total, these index cases caused 21 secondary cases from 12/23 (52%) households. Many households had 2-3 index cases as some of the family members had co-exposure outside the household. Among these 12 households, 4 households had 3 secondary cases, 1 household had two secondary cases, and 7 households had one secondary case. Within the 23 households with children, the secondary infection risk was 21/79 (26.6%) in Shenzhen.

We compared characteristics of contacts who became infected with those who did not (Table). No significant difference of age and sex was found between the secondary cases and non-cases. The secondary cases were more commonly to be immediate family (such as spouses, parents, siblings, and children) than non-cases (66.7% vs 37.9%, P<0.05). This could be associated with more intense contact than the extended family members. Most of the secondary cases lived together with the index cases, while only half for the non-cases (71.4% vs 51.7%, P<0.05) did so, and there was no secondary case among those not living with the index cases. Our findings are consistent with previous studies suggesting that closed environments with prolonged close contact may facilitate secondary transmission (5), and the transmission rate of infectious diseases within households was much greater than between individuals not living together (6). Thus, reducing close contact, keep contact distance including separated living environment within household, early quarantine of exposed household contacts and medical isolation of the ill people may be needed to reduce the onward transmission in household.

Interestingly, we found all secondary cases had contact with index cases during the period after symptom onset in the index cases, while just 67.2% of the non-cases had contact with them (P<0.05). Furthermore, the median duration of contact with post-illness-onset index cases was significantly longer among the secondary cases than non-cases (4.0 days, IQR: 2.0-5.0 vs 2.0, IQR: 0-4.0, P<0.05), although no significant difference was found for the median duration of contact with before-illness-onset index cases between the secondary cases and non-cases (2.0 days, IQR: 1.0-9.0 vs 5.0, IQR: 2.0-12.0, P=0.223) (Figure). This finding implied that the timing and duration of contact with the index cases was associated with the risk of secondary infection among household contacts. Recent studies also found the infectiousness was highest around the first day of illness onset and may even occur 1-2 days before illness onset (7, 8). Thus, early identification of the cases and diminishing the duration of contact, especially after the illness onset of the index cases, would be helpful to decrease the household transmission risk.

With recorded data on use of face mask among 11 households, we found that the secondary cases were much less likely to report wearing a mask at home compared to the non-cases (23.5% vs 75.0%, P<0.05). Our results support the recommendation that symptomatic individuals should use face mask as well as those who are caring for somebody with symptoms (9).

Unfortunately, we do not have information on other preventive measures, such as hand hygiene or household disinfection, which also could play a role in reducing household transmission. More studies are needed to evaluate the role of these factors in household transmission.

## Conclusions

Identifying risk and preventive factors for household transmission of SARS-CoV-2 is of great importance to guide preventive measures for onward transmission. The findings of this study suggested immediate family relationship, living together, and contact with index cases who had illness onset was associated higher odds of household transmission, while wearing face mask at home may be effective in reducing the risk of household transmission. A combination of different measures including active case-finding, medical isolation of confirmed and suspected cases, quarantine of close contacts and personal preventive measures such as wearing face mask were efficacious in the containment of household transmission in Shenzhen, China.

## Data Availability

All data in the manuscript was not publicly available now.

## Acknowledgments

We thank all staff from the Department of Public Health and the Department of Communicable Diseases Control and Prevention from Shenzhen Center for Disease Control and Prevention.

## Author Bio

Dr. Wei is an infectious disease epidemiologist at the Shenzhen Center for Disease Control and Prevention, Shenzhen, China. Her research interests are epidemiology and prevention and control of infectious disease.

---

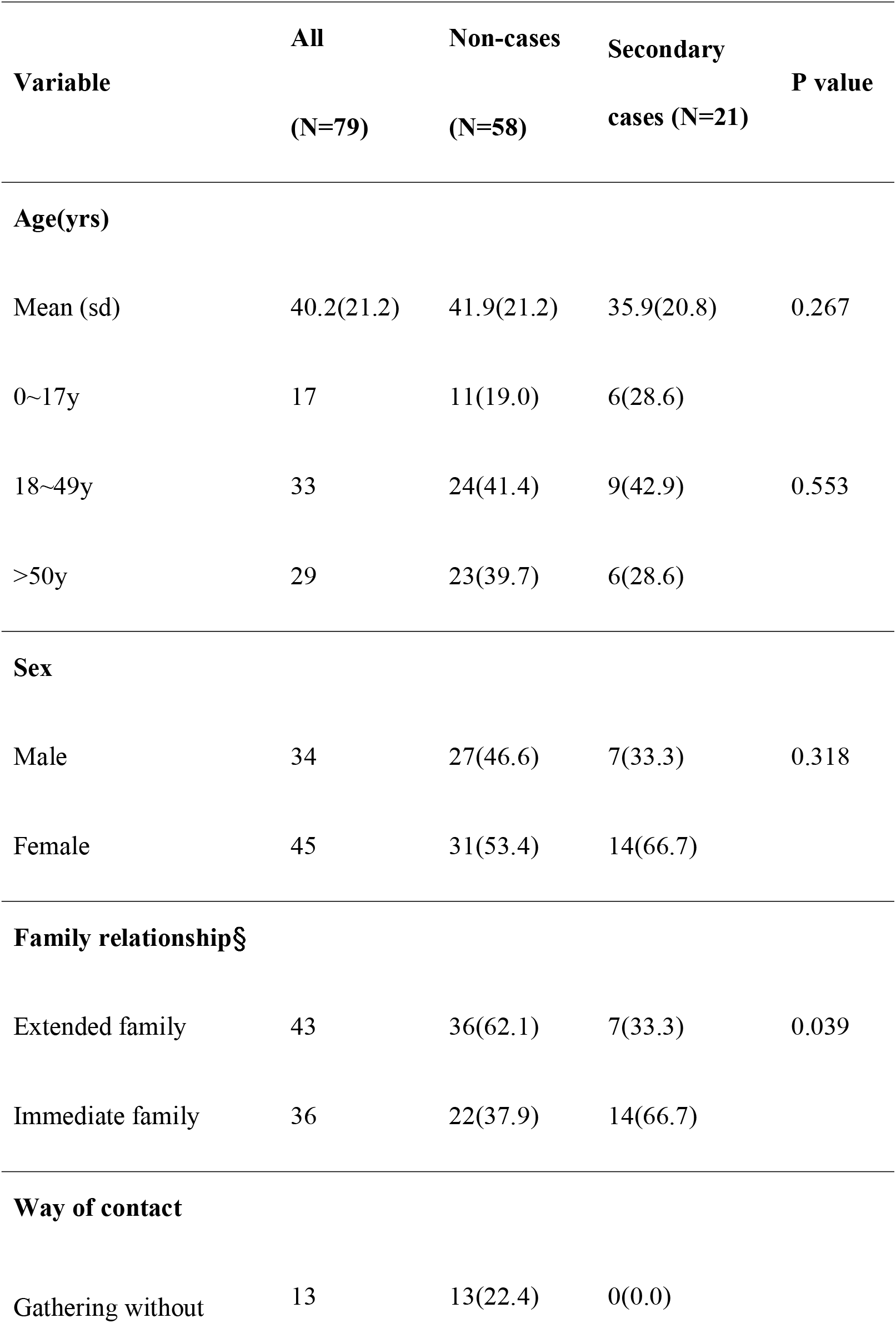
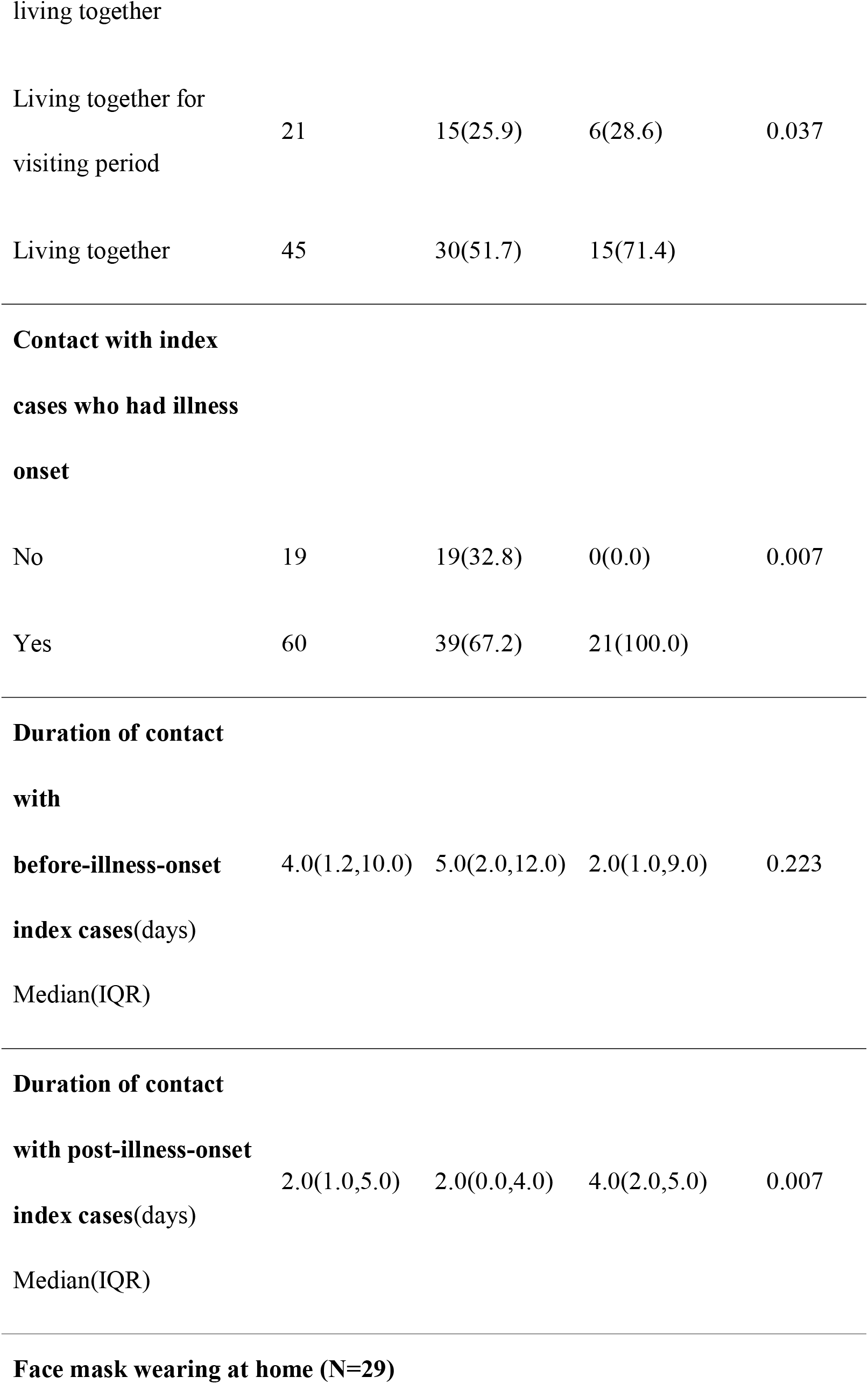
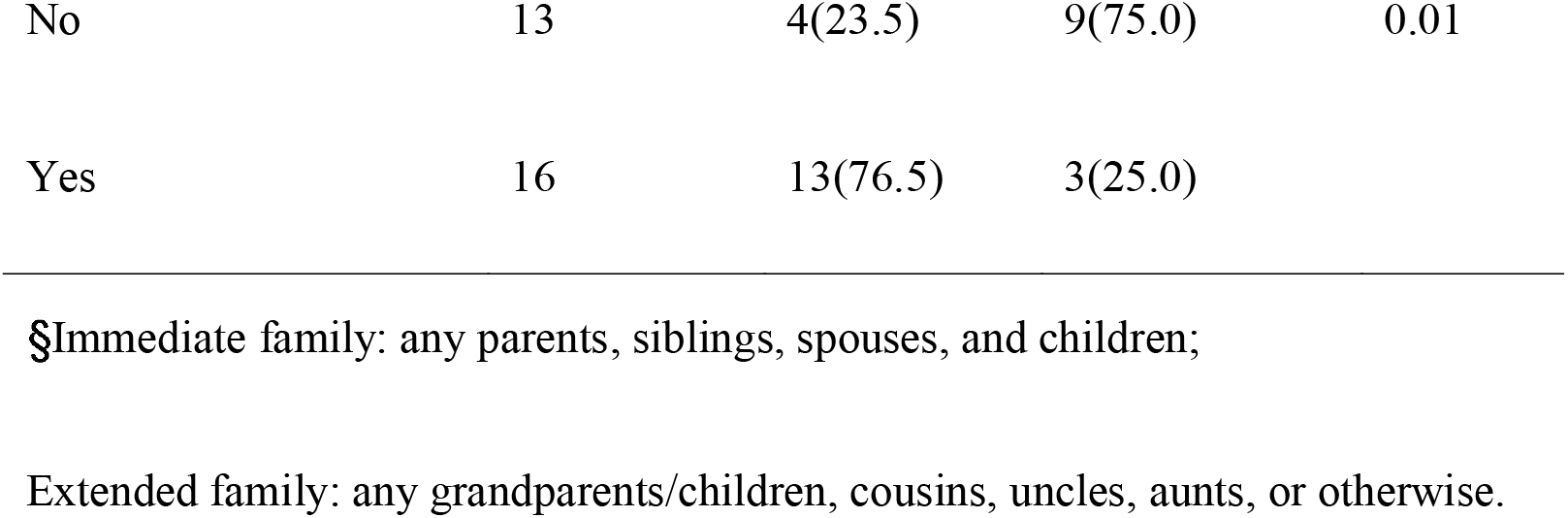
Table Characteristics of the secondary cases of COVID-19 and non-cases in households in Shenzhen, China.

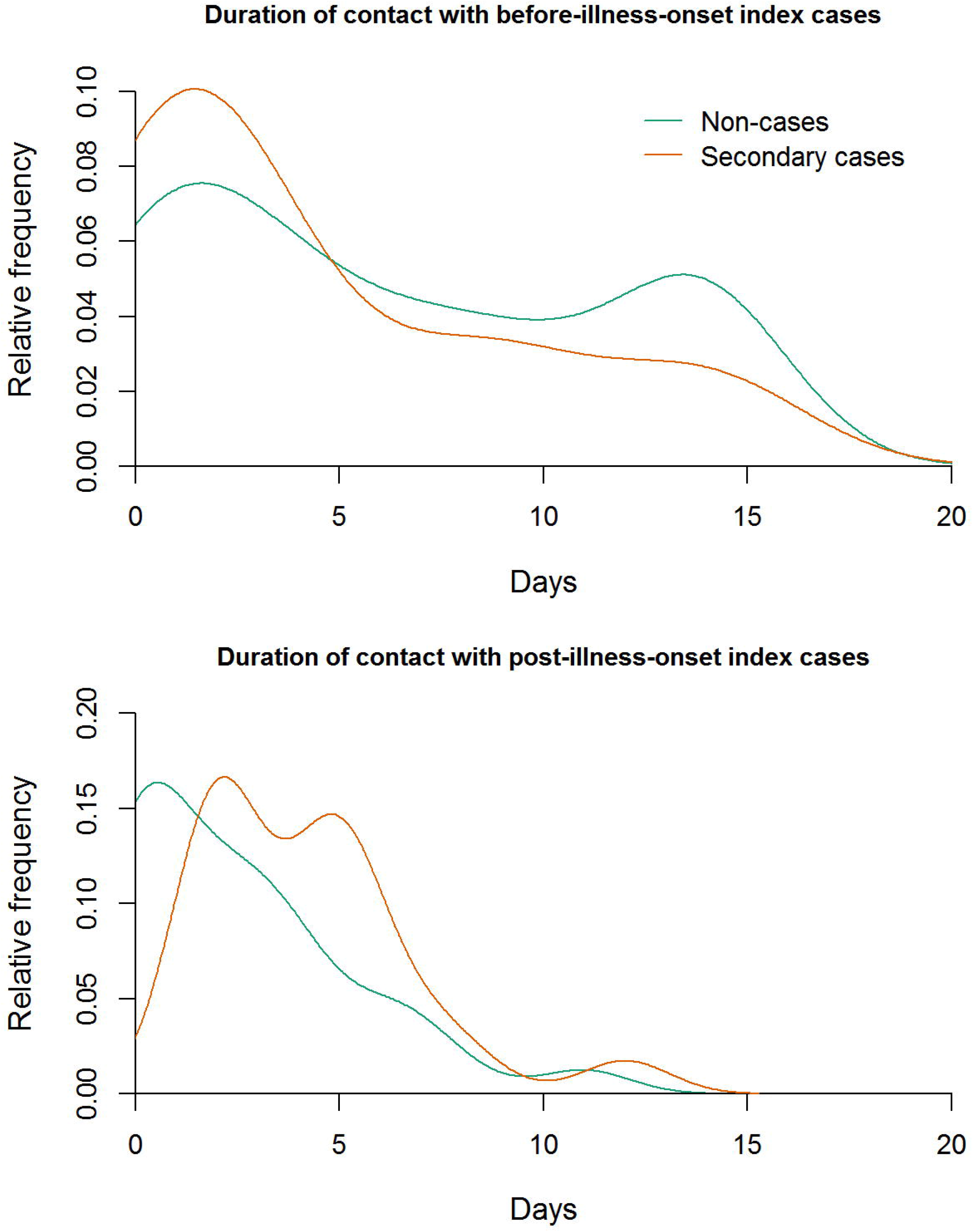
Figure Duration of the secondary household contact with the index case

## References

1. Coronavirus COVID-19 Global Cases by the Center for Systems Science and Engineering (CSSE) at Johns Hopkins University. 2020; Available from: https://coronavirus.jhu.edu/map.html.

2. Report of the WHO-China Joint Missionon Coronavirus Disease 2019 (COVID-19): World Health Organization; 2020.

3. Bi Q, Wu Y, Mei S, Ye C, Zou X, Zhang Z, et al. Epidemiology and transmission of COVID-19 in 391 cases and 1286 of their close contacts in Shenzhen, China: a retrospective cohort study. Lancet Infect Dis. In press 2020.

4. Wu Z, McGoogan JM. Characteristics of and Important Lessons From the Coronavirus Disease 2019 (COVID-19) Outbreak in China: Summary of a Report of 72314 Cases From the Chinese Center for Disease Control and Prevention. JAMA. 2020 Feb 24. doi: 10.1001/jama.2020.2648.

5. Nishiura H, Oshitani H, Kobayashi T, Saito T, Sunagawa T, Matsui T, et al. Closed environments facilitate secondary transmission of coronavirus disease 2019 (COVID-19). *medRxiv*. 2020; 2020.02.28.20029272.

6. Fraser C. Estimating individual and household reproduction numbers in an emerging epidemic. PLoS One. 2007; 2(8):e758-e.

7. Cowling BJ, Aiello A. Public health measures to slow community spread of COVID-19. J Infect Dis. 2020 Mar 20. doi: 10.1093/infdis/jiaa123.

8. Zou L, Ruan F, Huang M, Liang L, Huang H, Hong Z, et al. SARS-CoV-2 Viral Load in Upper Respiratory Specimens of Infected Patients. N Engl J Med. 2020; 382(12):1177–9.

9. Feng S, Shen C, Xia N, Song W, Fan M, Cowling BJ. Rational use of face masks in the COVID-19 pandemic. Lancet Respir Med. 2020 Mar 20. doi: 10.1016/S2213-2600(20)30134-X.

